# Metabolic Modulation of Glioblastoma by Dietary Intervention: A Systematic Review

**DOI:** 10.1101/2025.09.20.25336205

**Authors:** Maged T. Ghoche, Kenji Miki, Fanen Yuan, Megan Mantica, Sameer Agnihotri, Samuel K. McBrayer, Kalil G. Abdullah

**Author notes:** **Kalil G. Abdullah, MD, MSc, FAANS**, Associate Professor, Director, Translational Neuro Oncology, Department of Neurological Surgery, Hillman Cancer Center, University of Pittsburgh Medical Center, 5200 Centre Avenue, Suite 4B, Pittsburgh, PA 15232. **Author Contributions:** Conceptualization: KGA, MTG; Literature review and data collection: MTG, KM, FY; Data interpretation: KGA, MTG, SKM, SA, MM; Writing – original draft: KGA, MTG, KM, FY; Writing – review & editing: KGA, SA, SKM, MM, MTG; Supervision: KGA. **Funding:** No funding was received for the preparation of this manuscript. **Disclosures:** The authors have no personal, financial, or institutional interest in any of the drugs, materials, or devices described in this article.

## Abstract

**Background:** Glioblastoma (GBM) is characterized by aggressive clinical behavior and profound resistance to conventional therapies. As a result, non-pharmacologic strategies that exploit tumor metabolic vulnerabilities, particularly dietary interventions, are gaining attention. Ketogenic diets (KD) and methionine restriction (MR) aim to disrupt tumor bioenergetics and epigenetic regulation by modifying systemic nutrient availability.

**Objective:** To systematically evaluate preclinical and clinical evidence on the efficacy, safety, and mechanistic rationale of KD and MR in glioma treatment.

**Methods:** We conducted a systematic review of PubMed and Embase (through June 10, 2025), including preclinical and clinical studies involving KD or MR in gliomas. Eligible studies included in vivo preclinical models and clinical trials involving high grade gliomas. Data on study design, intervention characteristics, outcomes (survival, metabolic biomarkers, feasibility), and proposed mechanisms were extracted and synthesized narratively.

**Results:** A total of 31 studies were included: 10 preclinical and 13 clinical for KD; 6 preclinical and 2 clinical for MR. Preclinical studies demonstrated consistent anti-tumor effects, including survival prolongation and metabolic disruption, particularly when combined with radiotherapy or chemotherapy. Clinical outcomes were variable. KDs were feasible and well-tolerated, though adherence varied and survival benefits were inconsistent. MR showed promising metabolic effects and some clinical responses, but trials were limited by small sample sizes and high toxicity when combined with chemotherapy.

**Conclusion:** Dietary interventions such as KD and MR target critical metabolic vulnerabilities in gliomas and show strong preclinical rationale. However, clinical translation is challenged by tumor heterogeneity, metabolic adaptability, and dietary adherence. Future trials should incorporate metabolic imaging, flux analysis, and cross-disciplinary strategies to optimize and personalize dietary therapies as adjuncts in glioma care.

## Introduction

Glioblastoma (GBM) has an aggressive clinical course and profound resistance to existing treatments[1]. Two major impediments exist that have restricted GBM therapy development. The first is the impenetrability of the blood-brain barrier to most pharmaceutical agents[2]. Unlike peripheral tissues with unrestricted vascular access, GBMs residing within the brain parenchyma are largely shielded from systemic therapies by the blood–brain barrier, resulting in limited drug penetration and subtherapeutic exposure[3]. The second challenge is the vast heterogeneity among GBM. While dual checkpoint inhibition has successfully extended survival in some other cancers, GBM lacks these clonal hallmarks and has an intrinsic capacity to pivot from one oncogenic driver to another when targeted[4].

The limited efficacy of conventional therapies in altering the clinical course of GBM has catalyzed interest in systemically sustainable metabolic interventions capable of bypassing the blood–brain barrier. Among these, dietary strategies such as the ketogenic diet (KD) and methionine restriction (MR) have gained traction for their potential to reprogram the tumor microenvironment and are now under investigation in clinical trials[5], [6]. Both strategies aim to exploit the metabolic vulnerabilities of tumor cells, potentially enhancing therapeutic outcomes while minimizing systemic toxicity [5], [6].

This systematic review will the first to explore dietary interventions investigated in preclinical studies and those that have advanced to clinical trials, focusing on the KD and MR. We will evaluate their efficacy, safety profiles, mechanistic insights from preclinical models, and translational potential to complement existing therapies in treating gliomas and GBMs. By integrating preclinical findings with emerging clinical trial data, we evaluate the therapeutic potential of dietary interventions, identify critical gaps in current knowledge, and assess the feasibility of incorporating these strategies into contemporary neuro-oncology practice.

## Methods

### Search Strategy

We systematically searched PubMed and Embase for studies published before June 10, 2025, on dietary interventions in glioblastoma and other gliomas, including both preclinical and clinical research. Core search terms combined “glioblastoma” or “glioma” with “ketogenic diet,” “methionine restriction,” “dietary intervention,” or “amino acid restriction,” further refined with terms like “preclinical,” “clinical trial,” “IDH-mutant,” “IDH-wildtype,” “survival,” and “tumor suppression.” Search results were imported into a custom electronic database (Rayyan), with duplicates automatically removed. MTG, MK, and FY independently screened titles, abstracts, and full texts for eligibility. Disagreements were resolved by consensus or, if needed, by the senior author (KGA).

### Eligibility criteria

- We included preclinical and clinical studies evaluating dietary interventions in glioblastoma or other gliomas. Preclinical studies employed orthotopic or syngeneic glioma models, including both IDH-wildtype and IDH-mutant subtypes. Clinical studies included patients with high grade gliomas, primary or recurrent.
- Eligible interventions comprised various KD protocols, as well as MR via dietary or enzymatic methods (e.g., methionine lyase).
- Comparators included standard rodent diets for preclinical studies and standard-of-care therapy (e.g., temozolomide and radiotherapy) for clinical studies.
- Eligible designs included in vivo preclinical models, RCTs, single-arm trials, and prospective cohort studies. Excluded were in vitro studies, reviews, editorials, retrospective studies, and non-English publications.

### Data & Outcomes

Data were extracted detailing study design, population characteristics (glioma subtype, demographics, animal model), dietary intervention details (composition, duration, compliance), outcome metrics (survival, metabolic biomarkers, tumor volume), and risk of bias indicators (randomization, blinding, attrition).

The primary outcomes considered were overall survival (OS) and progression-free survival (PFS). Secondary outcomes included metabolic biomarkers, such as glucose-ketone index (GKI), β-hydroxybutyrate (βHB), and plasma methionine levels, tumor volume changes, feasibility (measured by adherence rates), and safety profiles, including the occurrence of adverse events.

Due to the heterogeneity in dietary protocols and study designs, a narrative synthesis was performed.

## Results

31 studies were included in the final synthesis (Fig.1). 10 preclinical[7], [8], [9], [10], [11], [12], [13], [14], [15], [16] and 13 clinical studies on KDs [14], [17], [18], [19], [20], [21], [22], [23], [24], [25], [26], [27], [28] and 6 preclinical [29], [30], [31], [32], [33], [34] and 2 clinical studies[35], [36] on MR. Figure 1 provides an overview of the screening process. Tables 1 and 2 summarize preclinical investigations on KDs and MR, respectively, outlining author and year, animal model, glioma model, intervention versus control conditions, outcome metrics, key findings, and proposed mechanisms of action. Table. 3 synthesizes clinical studies, reporting on study design, population, intervention, feasibility, safety, tolerability, effectiveness, and study limitations.

**Table.1.** Preclinical Studies of Ketogenic Diet (KD) and Caloric Restriction (CR) in Glioma Models.

| Author, year | Animal model | Glioma model | Intervention (vs. Control) | Outcome metrics | Key findings | Proposed mechanisms |
| --- | --- | --- | --- | --- | --- | --- |
| Abdelwahab et al., 2012 | C57BL/6 mice (♂, 10 wks) | GL261-luc2 orthotopic | KD: KetoCal® (4:1 fat:carb+protein) ± radiation ($2 \times 4$ Gy). Control: Standard diet (SD) ± radiation. | Survival, bioluminescence, blood βHB/glucose, histology. | - KD alone ↑ survival by 5 days (*p* < 0.005).<br>- KD + radiation induced tumor regression (9/11 mice; *p* < 0.0001).<br>- ↑ βHB, ↓ glucose (*p* < 0.0001).<br>- No recurrence post-diet. | KD sensitizes tumor to radiation via metabolic shift (↓ glucose, ↑ ketones), ROS, altered gene expression. |
| Seyfried et al., 2003 | C57BL/6J mice (♂, 10–12 wks) | CT-2A orthotopic | CR: SD or KD restricted to 40% intake. Control: SD/KD ad libitum. | Tumor weight, plasma glucose/βHB/IGF-1, body weight. | - CR ↓ tumor weight by 80% (*p* < 0.001).<br>- ↓ Glucose, ↑ βHB in CR groups.<br>- KD alone ineffective without CR. | CR exploits tumor glycolysis dependence on ketones ↓ inflammation. IGF-1 mediates anti-angiogenic effects. |
| **Maurer et al., 2011** | Athymic nude mice (♀, 8–10 wks) | LNT-229 xenograft | KD: High-fat (35.5%), low-carb (0.2%) ad libitum. Control: SD (55.6% carb). | Survival, blood glucose/βHB, MRI, histology (Ki-67), in vitro assays. | - KD ↑ βHB but no ↓ glucose/IGF-1 or tumor growth.<br>- No survival benefit (*p* = 0.288).<br>- Glioma cells unable to utilize βHB. | Glioma cells lack functional ketone metabolism. Non-C KD fails to reduce glucose. |
| **Zhou et al., 2007** | C57BL/6J & SCID mice (♂) | CT-2A, U87-MG orthotopic | KC-UR (KetoCal® ad libitum) vs. KC-R (30% CR). | Tumor weight, survival, plasma glucose/βHB, microvessel density, RT-PCR. | - KC-R ↓ tumor growth (65% CT-2A, 35% U87; *p* < 0.03).<br>- ↑ Survival (*p* < 0.01).<br>- ↓ Glucose, ↑ βHB (5–9×).<br>- ↓ Microvessel density (*p* < 0.05). | CR-KD exploits glucose dependency and defective keton metabolism (↓ β-OHBDH/SCOT). A angiogenic effects. |
| **Stafford et al., 2010** | C57BL/6 mice (♀) | GL261-luc orthotopic | KD: High-fat (78.8%), low-carb (8.36%). Control: SD. | Tumor bioluminescence, survival, ROS assays, gene expression, blood glucose/βHB. | - KD ↓ tumor growth (*p* < 0.05), ↑ survival (19d → 25d; *p* = 0.0196).<br>- ↓ ROS (*p* ≤ 0.001).<br>- Reversed tumor gene expression (e.g., ↓ COX2). | KD reduces ROS at shifts metabolism v gene expression changes (oxidative stress pathways). |
| **Javier et al., 2022** | Athymic nude & C57BL/6J mice | IDH1wt/mut gliomas | Cycling KD: Alternating weekly KD (90.5% fat) and SD. | Tumor volume, survival, blood glucose/ketones, in vitro proliferation. | - No survival/tumor growth differences (*p* > 0.05).<br>- KD ↑ ketones (*p* = 0.00022) but no glucose effect.<br>- IDH1mut tumors showed no KD vulnerability. | Unrestricted cycling KD ineffective. Tumors may adapt to ketones. |
| **Mukherjee et al., 2019** | VM/Dk & C57BL/6J mice (♂♀) | VM-M3/CT-2A orthotopic | KD-R (15–18% CR) ± glutamine antagonist (DON). | Bioluminescence, survival, histology, IHC (Ki-67/TNF-α), blood GKI, DON concentration. | - KD-R + DON ↓ tumor bioluminescence (75%; *p* < 0.0001).<br>- ↑ Survival (>40d vs. 15–18d controls).<br>- KD-R ↓ invasion, inflammation (↓ TNF-α/Iba-1).<br>- Enhanced DON delivery (*p* < 0.05). | Dual targeting of glycolysis (KD-R) & glutaminolysis (DON). KD-R exploits metabolic inflexibility; ketones neuroprotective. |
| **Ciusani et al., 2020** | C57BL/6N mice (♀) | GL261 orthotopic | KD-UR: High-fat (79.2%), low-carb (6%) ad libitum. Control: SD. | Survival, plasma glucose/βHB, MRI/MRS (metabolites), tumor volume. | - KD ↑ survival (*p* < 0.05).<br>- ↑ βHB in plasma/tumor (*p* < 0.025).<br>- ↓ GABA/NAA in tumor (*p* < 0.029). | KD-induced ketosis exploits tumor's limited ketone metabolism. Mild hypoglycemia stresses glucose-dependent tumors. |
| **Woolf et al., 2015** | C57BL/6 mice (♀) | GL261-Luc2 orthotopic | KD: KetoCal (4:1 ratio) ad libitum. Control: SD. | Blood βHB/glucose, bioluminescence, IHC (CD31/HIF-1α), MRI, WB (ZO-1/AQP4). | - KD ↑ βHB (*p* < 0.001), ↓ glucose (*p* < 0.01).<br>- ↓ Hypoxia (HIF-1α; *p* < 0.05), angiogenesis (CD31; *p* < 0.05).<br>- ↓ Edema, ↑ BBB integrity (ZO-1; *p* < 0.05). | KD reduces hypoxia-driven pathways (HIF-1α/NF-κB), normalizes vasculature, stabilizes BBB. Anti-angiogenic effects via VEGF/MMP downregulation. |
| **Rieger et al., 2014** | Athymic nude mice (♀) | U87MG orthotopic | KD ± bevacizumab (Bev). Control: SD ± Bev. | Survival, tumor volume (MRI), blood βHB, ATP/lactate/glucose. | - KD + Bev ↑ survival vs. Bev alone (58d vs. 52d; *p* < 0.05).<br>- Trend ↓ tumor volume/ATP. | KD exacerbates energy stress (↓ ATP) in tumors; synergizes with anti-angiogenic effects. |

**Table.2.**
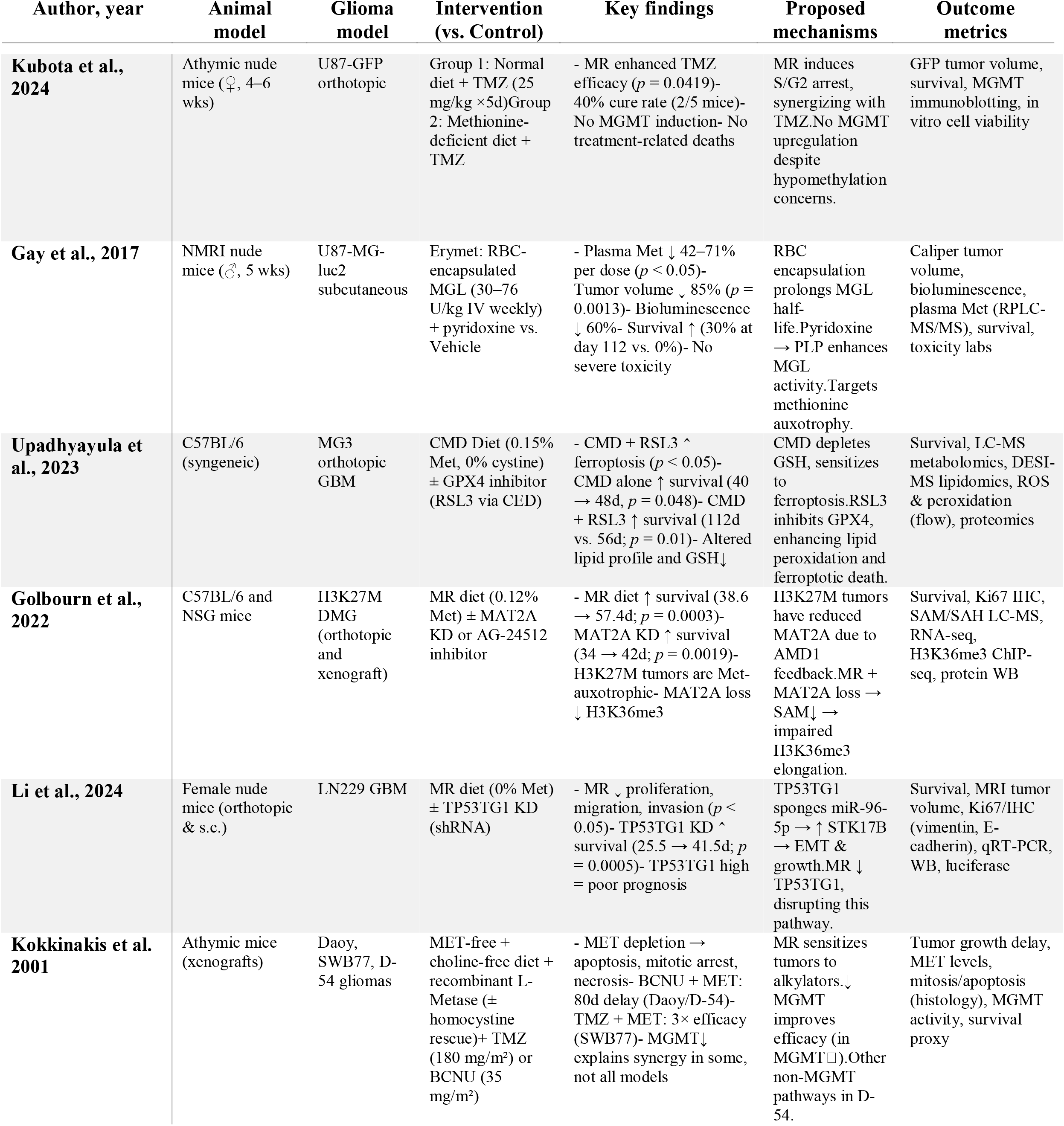
Preclinical Studies of Methionine Restriction in Glioma Models.

**Table 3.**
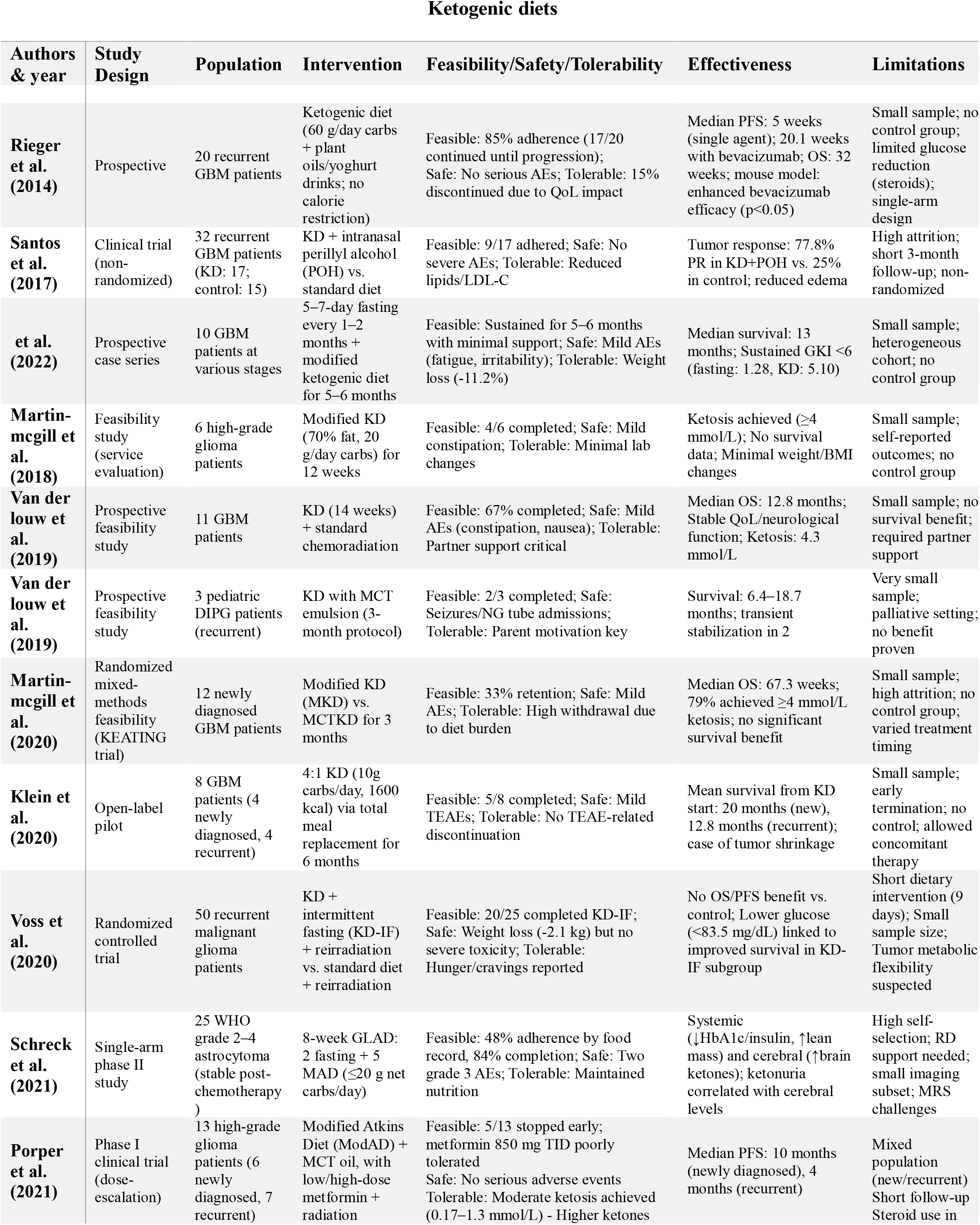
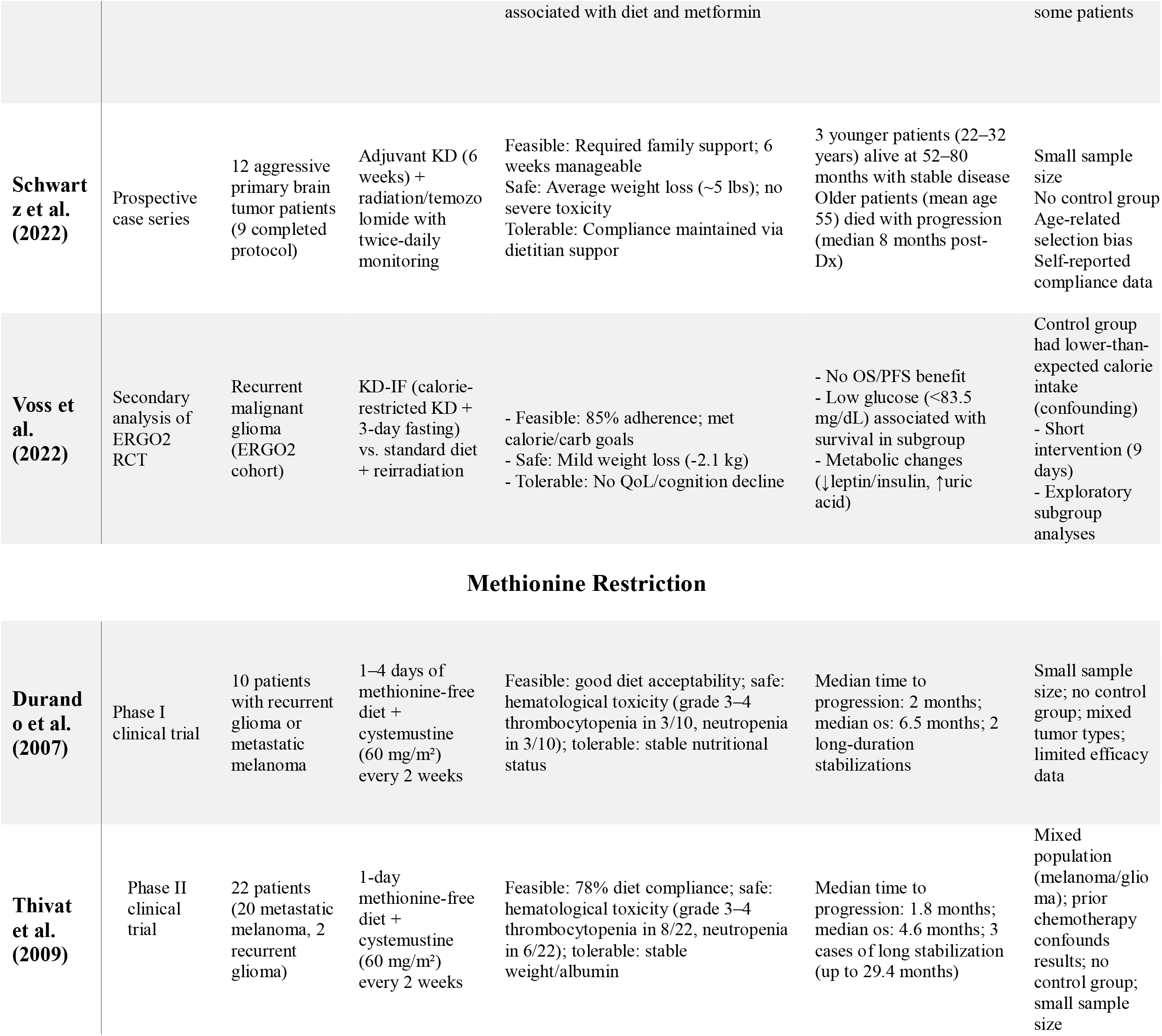
Summary of Clinical Studies Investigating Ketogenic and Methionine Restriction Diets in Glioma.

**Figure 1.**
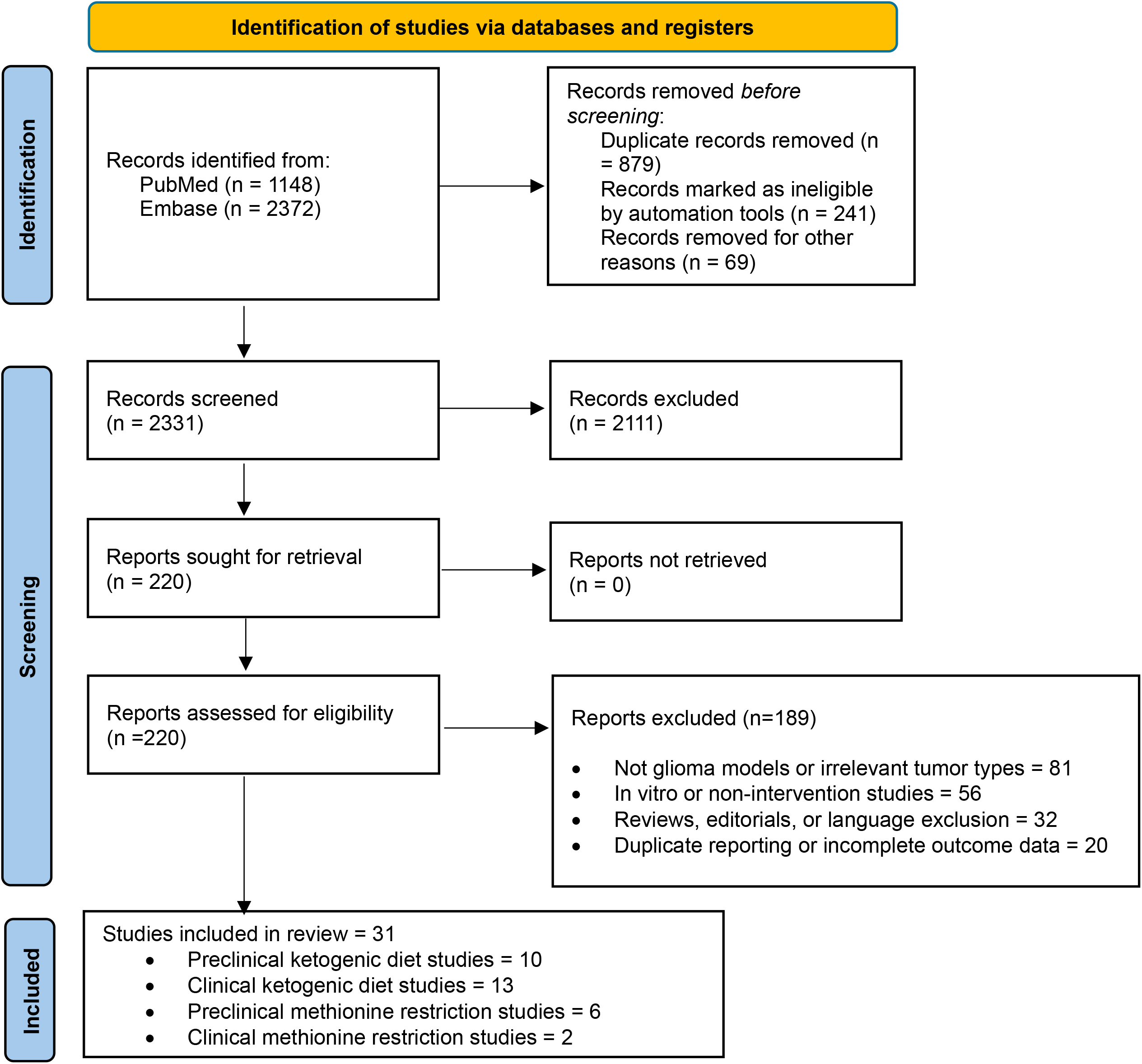
PRISMA flow diagram depicting title, abstract, and full-text screening in addition to reasons for study exclusion during full-text assessment.

### I The Ketogenic Diet

The KD is a non-pharmacologic strategy that targets glioma metabolism by exploiting tumor reliance on glucose. By restricting carbohydrates and increasing circulating ketone bodies, an energy source preferentially used by normal brain cells, KD creates a metabolic disadvantage for glycolytic tumor cells (Fig.2)[37]. KDs are high-fat, low-carbohydrate diets with adequate protein, shifting metabolism toward fat oxidation and ketogenesis[38]. Glucose deprivation reduces the insulin-to-glucagon ratio, diverting oxaloacetate toward gluconeogenesis and channeling acetyl-CoA into ketone production[39]. Many tumors, including gliomas, lack sufficient ketolytic enzymes, β-hydroxybutyrate dehydrogenase and succinyl-CoA:3-ketoacid CoA transferase, limiting their ability to utilize ketones for energy[40]. This creates a metabolic vulnerability where normal cells can adapt to ketosis, but glioma cells cannot [40].

Although KD variants continue to evolve, several main types are commonly used to improve adherence while preserving effectiveness[41]. The classic KD is highly restrictive, with 80–90% fat and strict 3:1 or 4:1 fat to carbohydrates + proteins ratios[42], [43]. The Medium-Chain Triglyceride (MCT) diet uses medium-chain fats for more carbs and protein [42], [43]. The Modified Atkins Diet (MAD) is more flexible, with looser ratios and no calorie or protein limits [42], [43]. The Low Glycemic Index Treatment (LGIT) allows more low-GI carbs, and the Mediterranean KD (MMKD) blends keto with Mediterranean dietary principles [42], [43].

#### 1. Preclinical Evidence on Ketogenic Therapy for Glioma

Preclinical studies report varied effects of KD in glioma models (Table.1). KD alone increases survival by 5–6 days (19d→25d, p=0.0196) and reduces tumor growth, associated with elevated βHB and decreased glucose (p<0.0001)[7], [10], [12], [13]. However, KD without CR shows limited or no significant anti-tumor benefit in some models[11], [41]. CR dramatically decreases tumor weight by up to 80% (p<0.001), reduces tumor growth by 65% (p<0.03), lowers glucose, and consistently enhances survival (p<0.01)[8], [16]. Combination strategies enhance KD efficacy; KD with radiation achieves tumor regression in 9/11 mice (p<0.0001), while KD with glutamine antagonist DON significantly prolongs survival (15–18d→>40d, p<0.0001) and reduces invasion and inflammation[8]. KD combined with bevacizumab extends survival (52d→58d, p<0.05), indicating synergy through reduced angiogenesis and tumor energy stress[41]. Overall, integrating KD or CR with complementary treatments provides the most promising outcomes, emphasizing the need for combined therapeutic strategies.

#### 2. Feasibility, Tolerability, and Patient Adherence

Feasibility of KD interventions in glioma patients was generally acceptable, with completion rates ranging from 33% to 85% (Table. 3). The highest feasibility was observed in one study, with 85% (17/20) of recurrent GBM patients completing the intervention[41], while another study reported the lowest, with only 33% retention in the KEATING feasibility trial[20].

Intermediate feasibility was seen in multiple studies; (67%, 11 adult GBM patients)[22], (84%, 25 astrocytoma patients)[27], and (63%, 5/8 completed a 6-month protocol)[23]. Pediatric studies also showed moderate feasibility (2/3 pediatric DIPG patients completed the 3-month protocol)[23], [28]. Factors enhancing feasibility included structured support (dietitians or family) and meal replacement strategies[23], [28]

Patient adherence was variable, with food-record-based adherence ranging from 48% to 85% (Table. 2)[14], [28]. Intermediate adherence was noted in ERGO2 where 85% of patients met calorie and carbohydrate goals in the cohort[25], and in other study where 53% (9/17) adhered to the KD+POH intervention[17]. The most significant barriers to adherence included diet complexity, side effects, and psychological burden, with high attrition rates reported multiple studies[17], [20], [26].

Tolerability was generally favorable. Across all studies, adverse events (AEs) were mostly mild to moderate, with no severe or life-threatening toxicities directly attributed to the diet. Common AEs included constipation, fatigue, nausea, irritability, and weight loss (up to 11.2% body weight loss[18]; average of ∼5 lbs[28]). The use of adjunct therapies such as metformin introduced some tolerability concerns; one study reported that 5/13 (38%) patients discontinued early due to poor tolerance of high-dose metformin[26]. Tolerability was significantly improved with strong partner or caregiver involvement[22], [28], structured meal replacement strategies, and close dietary monitoring^101^.

#### 3. Efficacy in Patients

Efficacy outcomes of KDs in glioma patients were highly heterogeneous (Table. 3). Median OS ranged from 8 months (older subgroup)[28] to 20 months (newly diagnosed patients)[23]. Intermediate OS values included 12.8 months[22], 13 months[18], and 67.3 weeks (∼15.6 months)[20]. In terms of PFS, a study reported a median of 5 weeks with KD alone and 20.1 weeks when combined with bevacizumab[14], while others reported 10 months (newly diagnosed) and 4 months (recurrent cases)[26]. In contrast, one study observed no formal PFS or OS outcomes but reported cerebral ketone elevation and systemic metabolic benefits[27].

In response outcomes, one study found that 77.8% (7/9) of KD+POH patients achieved partial response (PR) versus 25% (2/8) in the control group[17]. Others observed long-term survival (52–80 months) in 3 younger patients (22–32 years), suggesting a potential age-related benefit[28]. Metabolic markers like low glucose (<83.5 mg/dL) and sustained ketosis (≥4 mmol/L) were positively associated with survival in several studies[19], [24], [25]. However, no OS or PFS benefit was found in randomized or controlled indicating a lack of consistent evidence for clinical efficacy.

### II Targeting Methionine Metabolism

MR represents a promising strategy to exploit tumor-specific metabolic vulnerabilities in glioma. Most malignant cells, including GBM, exhibit methionine dependence, the inability to grow when methionine is replaced by homocysteine[26], [44]. This contrasts with normal cells, which retain flexibility through homocysteine remethylation [26], [44]. Methionine supports tumor growth via multiple pathways: protein synthesis, antioxidant defense through glutathione, and methylation reactions involving DNA, RNA, and histones[29], [45]. Methionine depletion disrupts these processes, triggering apoptosis, glutathione depletion, and MGMT suppression, thereby sensitizing tumors to alkylating agents such as temozolomide and BCNU[29], [46], [47].

In GBM, methionine dependence may arise from impaired homocysteine recycling, possibly driven by low methylcobalamin (vitamin B12) and decreased TCN2 transporter expression, particularly under hypoxia[48]. Tumor-initiating cells also show elevated MAT2A expression, leading to excessive methionine consumption and upregulated methionine cycling. These mechanisms contribute to subtype-specific vulnerabilities[47].

Moreover, glioblastoma’s high methionine uptake enables diagnostic imaging via ^11C-methionine PET, which offers superior sensitivity over FDG-PET in detecting tumor recurrence[49]. High methionine uptake is correlated with poor prognosis, reinforcing its clinical and metabolic significance [49].

#### 1. Preclinical Studies of Methionine Restriction in Glioma Models

Across six preclinical studies (Table. 2), MR strategies via diet, enzyme therapy, or genetic modulation demonstrated consistent tumor-suppressive and survival-prolonging effects in glioma models. Survival extension ranged from 6 to 56 days[31], [32], with MR alone increasing survival by 6–20 days, and MR combinations (e.g., with TMZ, RSL3, or MAT2A knockdown) extending survival by 16–56 days (p values all < 0.05) [31], [32]. Tumor volume reductions ranged from 60% to 85%, and tumor signal (e.g., bioluminescence, GFP) decreased by 60– 100%[32], [33], [34]. MR-induced cure rates were reported in 20–40% of mice102.

Mechanistically, MR consistently induced S/G2 arrest, apoptosis, necrosis, and mitotic delay, reduced GSH and SAM levels, and impaired epigenetic markers like H3K36me3[30], [31], [34] Ferroptosis was enhanced with MR+GPX4 inhibition, with corresponding changes in lipid peroxidation and ROS[31].MGMT suppression occurred variably, synergizing with TMZ in some models[32], [34], but not uniformly. MR downregulated pro-oncogenic lncRNAs (e.g., TP53TG1), altered EMT markers (vimentin↓, E-cadherin↑), and suppressed proliferation and migration by >50%[34].

Metabolically, plasma methionine was reduced by 42–71% per dose[33], and SAM suppression was confirmed via LC-MS[30]. No severe toxicities were reported across models.

#### 2. Clinical Trials of Methionine Restriction in Glioma

Clinical evidence on MR in glioma remains limited to two early-phase trials combining a short-term methionine-free diet with cystemustine chemotherapy (Table. 2). Across both studies[35], [36], MR was deemed feasible with diet compliance ranging from 72% to 78% and stable nutritional markers (e.g., weight, albumin). Hematological toxicity was the primary adverse event, with grade 3–4 thrombocytopenia occurring in 30–36% (3/10 to 8/22) and neutropenia in 30% (3/10 to 6/22) of patients. No treatment-related deaths were reported, and overall tolerability was acceptable.

Regarding efficacy, median time to progression ranged from 1.8 to 2 months, and median OS from 4.6 to 6.5 months. Both studies reported durable disease stabilization in select cases, with 2–3 patients experiencing long-term stability of >12 months, up to 29.4 months in one patient[36]. However, the clinical applicability of these findings is constrained by small sample sizes (n = 10–22), the inclusion of mixed tumor types (melanoma and glioma), and the lack of control arms.

## Discussion

Dietary interventions like KD and MR offer non-pharmacologic avenues to exploit metabolic dependencies in gliomas. Figure. 2 illustrates how this metabolic reprogramming reduces glucose and insulin levels, limits tumor cell survival pathways (e.g., PI3K/AKT), and induces apoptosis, especially when synergized with therapies. While preclinical evidence is strong, clinical outcomes remain inconsistent, limited by metabolic plasticity, variable adherence, and compensatory mechanisms such as MCT1 upregulation or glutamine anaplerosis.

**Figure 2.**
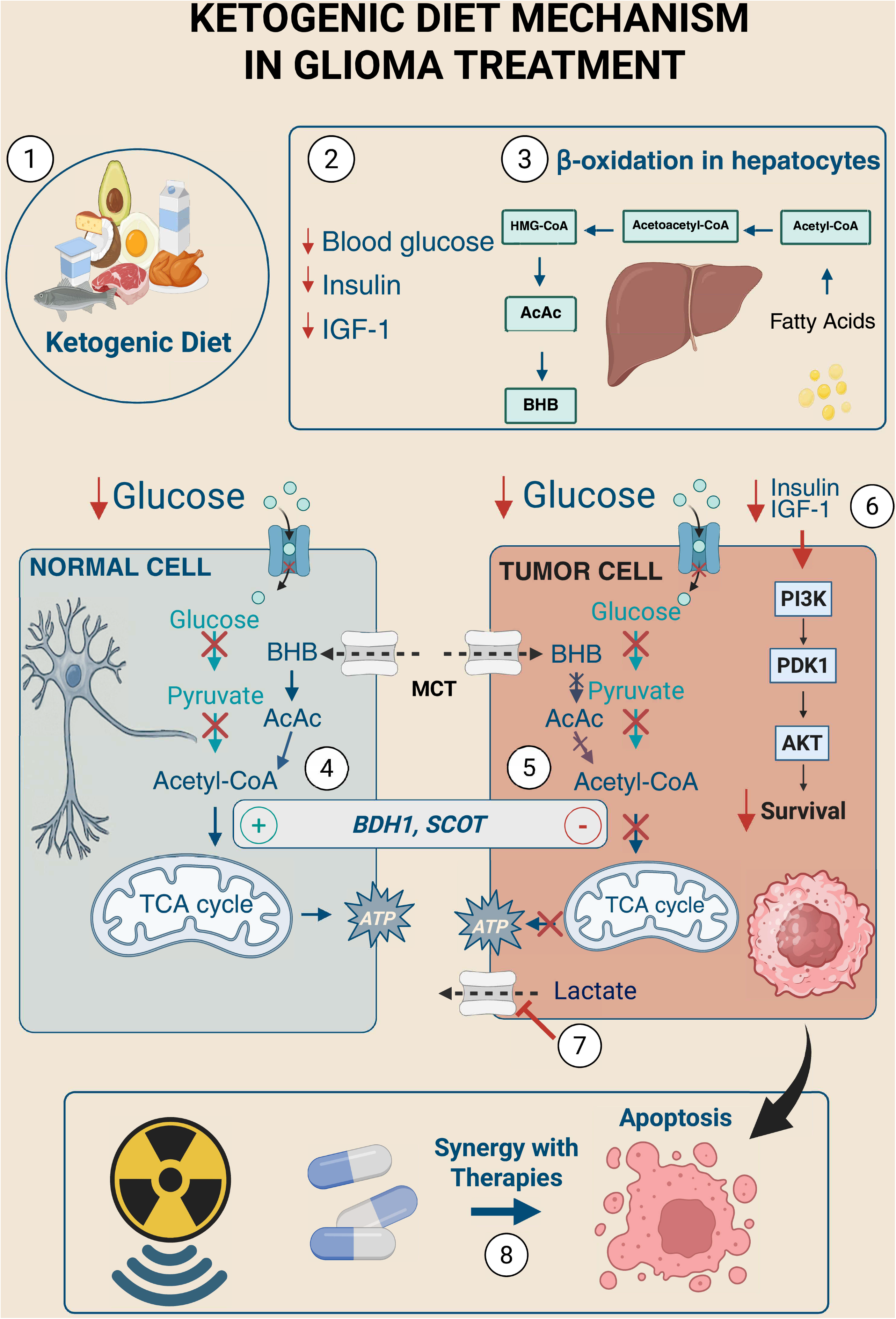
This schematic illustrates how the KD targets metabolic vulnerabilities in glioma cells while preserving normal brain cell function. (1) The KD is composed of high-fat, low-carbohydrate, and moderate-protein components that induce a metabolic shift away from glucose metabolism. (2) (3) In the liver, β-oxidation of fatty acids generates ketone bodies—primarily β-hydroxybutyrate (BHB) and acetoacetate (AcAc)—while reducing blood glucose, insulin, and insulin-like growth factor-1 (IGF-1) levels. (4) In normal brain cells, ketone bodies enter the mitochondria via monocarboxylate transporters (MCTs), are converted to acetyl-CoA, and fuel the tricarboxylic acid (TCA) cycle to produce ATP. These cells express sufficient levels of rate-limiting ketolytic enzymes including SCOT (succinyl-CoA:3-ketoacid CoA transferase) and BDH1 (β-hydroxybutyrate dehydrogenase). (5) In contrast, glioma cells demonstrate high glycolytic activity, depend on glucose, and often lack ketolytic enzymes. This renders them unable to effectively metabolize ketone bodies. (6) KD reduces systemic glucose and IGF-1, thereby downregulating oncogenic PI3K/Akt signaling and suppressing glycolysis and lactate production in glioma cells. (7) Ketone bodies compete for MCT1 on the tumor cell membrane, impairing lactate export. Trapped lactate in glycolytic tumor cells leads to intracellular acidosis and can inhibit further glycolysis – an unfavorable milieu for tumor survival. (8) KD also enhances tumor cell sensitivity to radiation and chemotherapy by lowering antioxidant defenses and reducing repair capacity.

MR diet limits intake by approximately 80% (to ∼2–3 mg/kg/day) and excludes most animal proteins and high-methionine plant foods (e.g., nuts, soy, lentils)[50]. Patients are guided toward low-methionine options like fruits, vegetables, and refined grains (Table. 4)[51]. To ensure adequate protein intake, MR diets often incorporate methionine-free medical foods. However, adherence remains a challenge due to poor taste and limited palatability[51], [52]. Newer approaches using protein-depleted or chemically modified foods may improve feasibility [51], [52]. These diets are typically isocaloric and require precise dietetic planning to maintain nutritional adequacy[52].

**Table 4.** Dietary Comparison Between the Ketogenic Diet and Methionine Restriction Diet.

| Category | Ketogenic diet | Methionine restriction diet |
| --- | --- | --- |
| Main goal | Severely restrict carbohydrates (typically <20–50 g/day) to induce ketosis; high fat, moderate protein. | Reduce methionine intake by ~80%; limit sulfur-containing amino acids while ensuring adequate energy and non-methionine protein sources. |
| Primary allowed foods | Fatty cuts of meat (beef, pork, lamb)<br>Fatty fish (salmon, mackerel, sardines) eggs<br>High-fat dairy (cheese, heavy cream, butter)<br>Leafy greens, non-starchy vegetables (broccoli, zucchini, cauliflower)<br>Nuts and seeds (in moderation)<br>Oils and fats (olive oil, coconut oil, mct oil) | Most fruits (apples, berries, bananas, citrus)<br>Most vegetables (including potatoes, carrots, beets)<br>Refined grains (white rice, white bread, pasta)<br>Low-protein breads and pastas<br>Low-methionine legumes (green peas, kidney beans, certain lentils)<br>Methionine-free medical foods (elemental amino acid formulas, specialized shakes) |
| Primary restricted foods | Grains (bread, pasta, rice, oats)<br>Starchy vegetables (potatoes, corn, peas)<br>Most fruits (due to sugar content)<br>Sugary foods (cakes, cookies, sodas, candy)<br>Legumes (beans, lentils, chickpeas)<br>Low-fat or fat-free dairy (due to carbs) | Meat (beef, pork, lamb)<br>Fish and seafood<br>Eggs- dairy products (milk, cheese, yogurt)<br>Soy products (tofu, soy milk, soy protein) lentils, peanuts, chickpeas (high methionine legumes)<br>Nuts and see-ds (due to methionine content) |

Cancer cells’ preference for aerobic glycolysis, known as the Warburg effect, has evolved from a byproduct of proliferation to a recognized driver of tumorigenesis[53]. In gliomas, this reprogramming is reinforced by alterations such as PI3K/Akt activation, c-MYC overexpression, p53 inactivation, and HIF-1α stabilization, promoting increased glucose and acetate metabolism through the TCA cycle and pentose phosphate pathway[53], [54], [55], [56], [57], [58], [59], [60].

IDH-mutant gliomas introduce further complexity through neomorphic mutations in IDH1/2, particularly IDH1 R132H, which convert α-ketoglutarate into the oncometabolite 2-hydroxyglutarate (2-HG)[61]. This metabolite impairs α-KG–dependent dioxygenases, leading to epigenetic repression and blocked neural differentiation[62], [63]. Vorasidenib, a blood-brain barrier-penetrant IDH1/2 inhibitor, has demonstrated promise by suppressing 2-HG production and exemplifies a new class of therapies targeting metabolic oncogenesis[64].

Beyond methionine, amino acid restriction strategies have revealed additional glioma vulnerabilities. Glioma cells upregulate serine synthesis (SSP) via ATF4 and c-MYC, diverting glycolytic intermediates toward nucleotide biosynthesis and redox maintenance[65]. Serine uptake is enhanced through SLC1A4/5 transporters, particularly in invasive regions[66]. Lysine restriction counters immune evasion mechanisms mediated by histone crotonylation, potentially enhancing response to MYC inhibitors or PD-1 blockade[67]. Glutamine dependency, especially pronounced in IDH1-mutant gliomas, supports biosynthesis and mTOR signaling[68], [69]. Glutamine-targeting agents like JHU-083 impair Cyclin D1 expression and prolong survival, likely by disrupting purine synthesis and mTOR integrity[68]. Arginine auxotrophy, observed in ASS1-silenced tumors, can be exploited via arginine-depleting agents such as ADI-PEG20, although this may suppress T-cell responses[68], [70]. Engineering ASS1-expressing CAR-T cells offers a strategy to mitigate this trade-off[71]. Tryptophan metabolism, through IDO1/TDO2 overexpression, promotes immunosuppression via kynurenine and AHR signaling[72], [73]. Despite the failure of IDO1 inhibitors in melanoma, combining them with checkpoint blockade or dietary interventions may hold promise in glioma[72].

## Conclusion

Broader targeting of amino acid metabolism underscores the therapeutic potential of dietary modulation but requires meticulous control due to rapid metabolic compensation. Successful translation demands a systems-level understanding of nutrient flux, metabolic shifts upon supplementation or withdrawal, and downstream effects on tumor progression and immunity. Window-of-opportunity or phase 0 trials, combined with radiolabel-based or microdialysis techniques, are essential for capturing dynamic metabolic changes in vivo. Interdisciplinary collaboration, spanning neurosurgery, oncology, nutrition, and pharmacology, will be critical to realizing metabolic interventions as viable adjuncts in glioma treatment.

## Data Availability

All data produced in the present study are available upon reasonable request to the authors

## References

[1] K. Urbańska, J. Sokołowska, M. Szmidt, and P. Sysa, “Glioblastoma multiforme - an overview.,” Contemp Oncol (Pozn), vol. 18, no. 5, pp. 307–12, 2014, doi: 10.5114/wo.2014.40559.

[2] O. Urhie et al., “Glioblastoma Survival Outcomes at a Tertiary Hospital in Appalachia: Factors Impacting the Survival of Patients Following Implementation of the Stupp Protocol.,” World Neurosurg, vol. 115, pp. e59–e66, Jul. 2018, doi: 10.1016/j.wneu.2018.03.163.

[3] Y. Zhao, L. Gan, L. Ren, Y. Lin, C. Ma, and X. Lin, “Factors influencing the blood-brain barrier permeability.,” Brain Res, vol. 1788, p. 147937, Aug. 2022, doi: 10.1016/j.brainres.2022.147937.

[4] A. Ghouzlani, S. Kandoussi, M. Tall, K. P. Reddy, S. Rafii, and A. Badou, “Immune Checkpoint Inhibitors in Human Glioma Microenvironment.,” Front Immunol, vol. 12, p. 679425, 2021, doi: 10.3389/fimmu.2021.679425.

[5] C. E. Champ et al., “Targeting metabolism with a ketogenic diet during the treatment of glioblastoma multiforme,” J Neurooncol, vol. 117, no. 1, pp. 125–131, Mar. 2014, doi: 10.1007/s11060-014-1362-0.

[6] R. Lakomy et al., “Real-World Evidence in Glioblastoma: Stupp’s Regimen After a Decade,” Front Oncol, vol. 10, Jul. 2020, doi: 10.3389/fonc.2020.00840.

[7] M. G. Abdelwahab et al., “The Ketogenic Diet Is an Effective Adjuvant to Radiation Therapy for the Treatment of Malignant Glioma,” PLoS One, vol. 7, no. 5, p. e36197, May 2012, doi: 10.1371/journal.pone.0036197.

[8] P. Mukherjee, L. E. Abate, and T. N. Seyfried, “Antiangiogenic and Proapoptotic Effects of Dietary Restriction on Experimental Mouse and Human Brain Tumors,” Clinical Cancer Research, vol. 10, no. 16, pp. 5622–5629, Aug. 2004, doi: 10.1158/1078-0432.CCR-04-0308.

[9] G. D. Maurer et al., “Differential utilization of ketone bodies by neurons and glioma cell lines: a rationale for ketogenic diet as experimental glioma therapy,” BMC Cancer, vol. 11, no. 1, p. 315, Dec. 2011, doi: 10.1186/1471-2407-11-315.

[10] P. Stafford, M. G. Abdelwahab, D. Y. Kim, M. C. Preul, J. M. Rho, and A. C. Scheck, “The ketogenic diet reverses gene expression patterns and reduces reactive oxygen species levels when used as an adjuvant therapy for glioma,” Nutr Metab (Lond), vol. 7, no. 1, p. 74, Dec. 2010, doi: 10.1186/1743-7075-7-74.

[11] R. Javier, W. Wang, M. Drumm, K. McCortney, J. N. Sarkaria, and C. Horbinski, “The efficacy of an unrestricted cycling ketogenic diet in preclinical models of IDH wild-type and IDH mutant glioma,” PLoS One, vol. 17, no. 2, p. e0257725, Feb. 2022, doi: 10.1371/journal.pone.0257725.

[12] E. Ciusani et al., “MR-Spectroscopy and Survival in Mice with High Grade Glioma Undergoing Unrestricted Ketogenic Diet,” Nutr Cancer, vol. 73, no. 11–12, pp. 2315–2322, Dec. 2021, doi: 10.1080/01635581.2020.1822423.

[13] E. C. Woolf et al., “The Ketogenic Diet Alters the Hypoxic Response and Affects Expression of Proteins Associated with Angiogenesis, Invasive Potential and Vascular Permeability in a Mouse Glioma Model,” PLoS One, vol. 10, no. 6, p. e0130357, Jun. 2015, doi: 10.1371/journal.pone.0130357.

[14] J. Rieger et al., “ERGO: A pilot study of ketogenic diet in recurrent glioblastoma,” Int J Oncol, vol. 44, no. 6, pp. 1843–1852, Jun. 2014, doi: 10.3892/ijo.2014.2382.

[15] P. Mukherjee et al., “Therapeutic benefit of combining calorie-restricted ketogenic diet and glutamine targeting in late-stage experimental glioblastoma,” Commun Biol, vol. 2, no. 1, p. 200, May 2019, doi: 10.1038/s42003-019-0455-x.

[16] W. Zhou, P. Mukherjee, M. A. Kiebish, W. T. Markis, J. G. Mantis, and T. N. Seyfried, “The calorically restricted ketogenic diet, an effective alternative therapy for malignant brain cancer,” Nutr Metab (Lond), vol. 4, no. 1, p. 5, Dec. 2007, doi: 10.1186/1743-7075-4-5.

[17] J. Santos et al., “Efficacy of a ketogenic diet with concomitant intranasal perillyl alcohol as a novel strategy for the therapy of recurrent glioblastoma,” Oncol Lett, Nov. 2017, doi: 10.3892/ol.2017.7362.

[18] M. C. L. Phillips et al., “Feasibility and Safety of a Combined Metabolic Strategy in Glioblastoma Multiforme: A Prospective Case Series,” J Oncol, vol. 2022, pp. 1–10, Oct. 2022, doi: 10.1155/2022/4496734.

[19] K. J. Martin-McGill, A. G. Marson, C. Tudur Smith, and M. D. Jenkinson, “The Modified Ketogenic Diet in Adults with Glioblastoma: An Evaluation of Feasibility and Deliverability within the National Health Service,” Nutr Cancer, vol. 70, no. 4, pp. 643–649, May 2018, doi: 10.1080/01635581.2018.1460677.

[20] K. J. Martin-McGill et al., “Ketogenic diets as an adjuvant therapy for glioblastoma (KEATING): a randomized, mixed methods, feasibility study,” J Neurooncol, vol. 147, no. 1, pp. 213–227, Mar. 2020, doi: 10.1007/s11060-020-03417-8.

[21] E. J. T. M. van der Louw, R. E. Reddingius, J. F. Olieman, R. F. Neuteboom, and C. E. Catsman-Berrevoets, “Ketogenic diet treatment in recurrent diffuse intrinsic pontine glioma in children: A safety and feasibility study,” Pediatr Blood Cancer, vol. 66, no. 3, Mar. 2019, doi: 10.1002/pbc.27561.

[22] E. J. T. M. van der Louw et al., “Ketogenic diet treatment as adjuvant to standard treatment of glioblastoma multiforme: a feasibility and safety study,” Ther Adv Med Oncol, vol. 11, Jan. 2019, doi: 10.1177/1758835919853958.

[23] P. Klein, I. Tyrlikova, G. Zuccoli, A. Tyrlik, and J. C. Maroon, “Treatment of glioblastoma multiforme with ‘classic’ 4:1 ketogenic diet total meal replacement,” Cancer Metab, vol. 8, no. 1, p. 24, Dec. 2020, doi: 10.1186/s40170-020-00230-9.

[24] M. Voss et al., “Short-term fasting in glioma patients: analysis of diet diaries and metabolic parameters of the ERGO2 trial,” Eur J Nutr, vol. 61, no. 1, pp. 477–487, Feb. 2022, doi: 10.1007/s00394-021-02666-1.

[25] M. Voss et al., “Short-term fasting in glioma patients: analysis of diet diaries and metabolic parameters of the ERGO2 trial,” Eur J Nutr, vol. 61, no. 1, pp. 477–487, Feb. 2022, doi: 10.1007/s00394-021-02666-1.

[26] K. Porper et al., “A Phase I clinical trial of dose-escalated metabolic therapy combined with concomitant radiation therapy in high-grade glioma,” J Neurooncol, vol. 153, no. 3, pp. 487–496, Jul. 2021, doi: 10.1007/s11060-021-03786-8.

[27] K. C. Schreck et al., “Feasibility and Biological Activity of a Ketogenic/Intermittent-Fasting Diet in Patients With Glioma,” Neurology, vol. 97, no. 9, Aug. 2021, doi: 10.1212/WNL.0000000000012386.

[28] K. A. Schwartz et al., “Long Term Survivals in Aggressive Primary Brain Malignancies Treated With an Adjuvant Ketogenic Diet,” Front Nutr, vol. 9, May 2022, doi: 10.3389/fnut.2022.770796.

[29] D. M. Kokkinakis et al., “Synergy between methionine stress and chemotherapy in the treatment of brain tumor xenografts in athymic mice.,” Cancer Res, vol. 61, no. 10, pp. 4017–23, May 2001.

[30] B. J. Golbourn et al., “Loss of MAT2A compromises methionine metabolism and represents a vulnerability in H3K27M mutant glioma by modulating the epigenome,” Nat Cancer, vol. 3, no. 5, pp. 629–648, Apr. 2022, doi: 10.1038/s43018-022-00348-3.

[31] P. S. Upadhyayula et al., “Dietary restriction of cysteine and methionine sensitizes gliomas to ferroptosis and induces alterations in energetic metabolism,” Nat Commun, vol. 14, no. 1, p. 1187, Mar. 2023, doi: 10.1038/s41467-023-36630-w.

[32] Y. Kubota et al., “Methionine restriction of glioma does not induce MGMT and greatly improves temozolomide efficacy in an orthotopic nude-mouse model: A potential curable approach to a clinically-incurable disease,” Biochem Biophys Res Commun, vol. 695, p. 149418, Feb. 2024, doi: 10.1016/j.bbrc.2023.149418.

[33] F. Gay et al., “Methionine tumor starvation by erythrocyte-encapsulated methionine gamma-lyase activity controlled with per os vitamin B6,” Cancer Med, vol. 6, no. 6, pp. 1437–1452, Jun. 2017, doi: 10.1002/cam4.1086.

[34] J. Li et al., “Methionine deprivation inhibits glioma proliferation and EMT via the TP53TG1/miR-96-5p/STK17B ceRNA pathway,” NPJ Precis Oncol, vol. 8, no. 1, p. 270, Nov. 2024, doi: 10.1038/s41698-024-00763-y.

[35] X. Durando et al., “Optimal Methionine-Free Diet Duration for Nitrourea Treatment: A Phase I Clinical Trial,” Nutr Cancer, vol. 60, no. 1, pp. 23–30, Dec. 2007, doi: 10.1080/01635580701525877.

[36] E. Thivat et al., “Phase II trial of the association of a methionine-free diet with cystemustine therapy in melanoma and glioma.,” Anticancer Res, vol. 29, no. 12, pp. 5235–40, Dec. 2009.

[37] R. L. Veech, “Ketone ester effects on metabolism and transcription,” J Lipid Res, vol. 55, no. 10, pp. 2004–2006, Oct. 2014, doi: 10.1194/jlr.R046292.

[38] A. L. Hartman and E. P. G. Vining, “Clinical Aspects of the Ketogenic Diet,” Epilepsia, vol. 48, no. 1, pp. 31–42, Jan. 2007, doi: 10.1111/j.1528-1167.2007.00914.x.

[39] J. C. Newman and E. Verdin, “β-Hydroxybutyrate: A Signaling Metabolite,” Annu Rev Nutr, vol. 37, no. 1, pp. 51–76, Aug. 2017, doi: 10.1146/annurev-nutr-071816-064916.

[40] M. J. Tisdale, “Role of acetoacetyl-CoA synthetase in acetoacetate utilization by tumor cells.,” Cancer Biochem Biophys, vol. 7, no. 2, pp. 101–7, Jun. 1984.

[41] G. D. Maurer et al., “Differential utilization of ketone bodies by neurons and glioma cell lines: a rationale for ketogenic diet as experimental glioma therapy,” BMC Cancer, vol. 11, no. 1, p. 315, Dec. 2011, doi: 10.1186/1471-2407-11-315.

[42] D. Malinowska and M. Żendzian-Piotrowska, “Ketogenic Diet: A Review of Composition Diversity, Mechanism of Action and Clinical Application,” J Nutr Metab, vol. 2024, no. 1, Jan. 2024, doi: 10.1155/2024/6666171.

[43] M. Barzegar, M. Afghan, V. Tarmahi, M. Behtari, S. Rahimi Khamaneh, and S. Raeisi, “Ketogenic diet: overview, types, and possible anti-seizure mechanisms,” Nutr Neurosci, vol. 24, no. 4, pp. 307–316, Apr. 2021, doi: 10.1080/1028415X.2019.1627769.

[44] M. E. Anderson, “Glutathione: an overview of biosynthesis and modulation,” Chem Biol Interact, vol. 111–112, pp. 1–14, Apr. 1998, doi: 10.1016/S0009-2797(97)00146-4.

[45] T. Thomas* and T. J. Thomas, “Polyamines in cell growth and cell death: molecular mechanisms and therapeutic applications,” Cellular and Molecular Life Sciences, vol. 58, no. 2, pp. 244–258, Feb. 2001, doi: 10.1007/PL00000852.

[46] T. Fiskerstrand, B. Christensen, O. B. Tysnes, P. M. Ueland, and H. Refsum, “Development and reversion of methionine dependence in a human glioma cell line: relation to homocysteine remethylation and cobalamin status.,” Cancer Res, vol. 54, no. 18, pp. 4899–906, Sep. 1994.

[47] M. L. Sowers and L. C. Sowers, “Glioblastoma and Methionine Addiction,” Int J Mol Sci, vol. 23, no. 13, p. 7156, Jun. 2022, doi: 10.3390/ijms23137156.

[48] K. Zhang et al., “Proteome Analysis of Hypoxic Glioblastoma Cells Reveals Sequential Metabolic Adaptation of One-Carbon Metabolic Pathways,” Molecular & Cellular Proteomics, vol. 16, no. 11, pp. 1906–1921, Nov. 2017, doi: 10.1074/mcp.RA117.000154.

[49] K. Nakajo et al., “Maximum 11C-methionine PET uptake as a prognostic imaging biomarker for newly diagnosed and untreated astrocytic glioma,” Sci Rep, vol. 12, no. 1, p. 546, Jan. 2022, doi: 10.1038/s41598-021-04216-5.

[50] A. Elorza et al., “HIF2α Acts as an mTORC1 Activator through the Amino Acid Carrier SLC7A5,” Mol Cell, vol. 48, no. 5, pp. 681–691, Dec. 2012, doi: 10.1016/j.molcel.2012.09.017.

[51] B. E. Hasek et al., “Dietary methionine restriction enhances metabolic flexibility and increases uncoupled respiration in both fed and fasted states,” American Journal of Physiology-Regulatory, Integrative and Comparative Physiology, vol. 299, no. 3, pp. R728–R739, Sep. 2010, doi: 10.1152/ajpregu.00837.2009.

[52] H. Fang, K. P. Stone, D. Wanders, L. A. Forney, and T. W. Gettys, “The Origins, Evolution, and Future of Dietary Methionine Restriction,” Annu Rev Nutr, vol. 42, no. 1, pp. 201–226, Aug. 2022, doi: 10.1146/annurev-nutr-062320-111849.

[53] O. Warburg, F. Wind, and E. Negelein, “THE METABOLISM OF TUMORS IN THE BODY.,” J Gen Physiol, vol. 8, no. 6, pp. 519–30, Mar. 1927, doi: 10.1085/jgp.8.6.519.

[54] T. Mashimo et al., “Acetate Is a Bioenergetic Substrate for Human Glioblastoma and Brain Metastases,” Cell, vol. 159, no. 7, pp. 1603–1614, Dec. 2014, doi: 10.1016/j.cell.2014.11.025.

[55] I. Marin-Valencia et al., “Analysis of Tumor Metabolism Reveals Mitochondrial Glucose Oxidation in Genetically Diverse Human Glioblastomas in the Mouse Brain In Vivo,” Cell Metab, vol. 15, no. 6, pp. 827–837, Jun. 2012, doi: 10.1016/j.cmet.2012.05.001.

[56] I. Papandreou, R. A. Cairns, L. Fontana, A. L. Lim, and N. C. Denko, “HIF-1 mediates adaptation to hypoxia by actively downregulating mitochondrial oxygen consumption,” Cell Metab, vol. 3, no. 3, pp. 187–197, Mar. 2006, doi: 10.1016/j.cmet.2006.01.012.

[57] N. C. Denko, “Hypoxia, HIF1 and glucose metabolism in the solid tumour,” Nat Rev Cancer, vol. 8, no. 9, pp. 705–713, Sep. 2008, doi: 10.1038/nrc2468.

[58] C. Wanka, J. P. Steinbach, and J. Rieger, “Tp53-induced Glycolysis and Apoptosis Regulator (TIGAR) Protects Glioma Cells from Starvation-induced Cell Death by Up-regulating Respiration and Improving Cellular Redox Homeostasis,” Journal of Biological Chemistry, vol. 287, no. 40, pp. 33436–33446, Sep. 2012, doi: 10.1074/jbc.M112.384578.

[59] R. L. Elstrom et al., “Akt Stimulates Aerobic Glycolysis in Cancer Cells,” Cancer Res, vol. 64, no. 11, pp. 3892–3899, Jun. 2004, doi: 10.1158/0008-5472.CAN-03-2904.

[60] R. B. Robey and N. Hay, “Is Akt the ‘Warburg kinase’?—Akt-energy metabolism interactions and oncogenesis,” Semin Cancer Biol, vol. 19, no. 1, pp. 25–31, Feb. 2009, doi: 10.1016/j.semcancer.2008.11.010.

[61] L. Evans et al., “IDH-mutant gliomas in children and adolescents - from biology to clinical trials,” Front Oncol, vol. 14, Jan. 2025, doi: 10.3389/fonc.2024.1515538.

[62] I. K. Mellinghoff et al., “Vorasidenib, a Dual Inhibitor of Mutant IDH1/2, in Recurrent or Progressive Glioma; Results of a First-in-Human Phase I Trial,” Clinical Cancer Research, vol. 27, no. 16, pp. 4491–4499, Aug. 2021, doi: 10.1158/1078-0432.CCR-21-0611.

[63] N. Sharma et al., “Isocitrate dehydrogenase mutations in gliomas: A review of current understanding and trials,” Neurooncol Adv, vol. 5, no. 1, Jan. 2023, doi: 10.1093/noajnl/vdad053.

[64] D. D. Shi et al., “De novo pyrimidine synthesis is a targetable vulnerability in IDH mutant glioma,” Cancer Cell, vol. 40, no. 9, pp. 939-956.e16, Sep. 2022, doi: 10.1016/j.ccell.2022.07.011.

[65] S. Pan, M. Fan, Z. Liu, X. Li, and H. Wang, “Serine, glycine and one-carbon metabolism in cancer (Review),” Int J Oncol, vol. 58, no. 2, pp. 158–170, Dec. 2020, doi: 10.3892/ijo.2020.5158.

[66] A. J. Scott et al., “Rewiring of cortical glucose metabolism fuels human brain cancer growth,” Oct. 25, 2023. doi: 10.1101/2023.10.24.23297489.

[67] J. Márquez, F. J. Alonso, J. M. Matés, J. A. Segura, M. Martín-Rufián, and J. A. Campos-Sandoval, “Glutamine Addiction In Gliomas,” Neurochem Res, vol. 42, no. 6, pp. 1735–1746, Jun. 2017, doi: 10.1007/s11064-017-2212-1.

[68] X. Hou, S. Chen, P. Zhang, D. Guo, and B. Wang, “Targeted Arginine Metabolism Therapy: A Dilemma in Glioma Treatment,” Front Oncol, vol. 12, Jul. 2022, doi: 10.3389/fonc.2022.938847.

[69] A. S. Yamashita, M. da Costa Rosa, V. Stumpo, R. Rais, B. S. Slusher, and G. J. Riggins, “The glutamine antagonist prodrug JHU-083 slows malignant glioma growth and disrupts mTOR signaling,” Neurooncol Adv, vol. 3, no. 1, Jan. 2021, doi: 10.1093/noajnl/vdaa149.

[70] L. Fultang et al., “Metabolic engineering against the arginine microenvironment enhances CAR-T cell proliferation and therapeutic activity,” Blood, vol. 136, no. 10, pp. 1155–1160, Sep. 2020, doi: 10.1182/blood.2019004500.

[71] M. Platten, E. A. A. Nollen, U. F. Röhrig, F. Fallarino, and C. A. Opitz, “Tryptophan metabolism as a common therapeutic target in cancer, neurodegeneration and beyond,” Nat Rev Drug Discov, vol. 18, no. 5, pp. 379–401, May 2019, doi: 10.1038/s41573-019-0016-5.

[72] G. V Long et al., “Epacadostat plus pembrolizumab versus placebo plus pembrolizumab in patients with unresectable or metastatic melanoma (ECHO-301/KEYNOTE-252): a phase 3, randomised, double-blind study,” Lancet Oncol, vol. 20, no. 8, pp. 1083–1097, Aug. 2019, doi: 10.1016/S1470-2045(19)30274-8.

[73] I. Cervenka, L. Z. Agudelo, and J. L. Ruas, “Kynurenines: Tryptophan’s metabolites in exercise, inflammation, and mental health,” Science (1979), vol. 357, no. 6349, Jul. 2017, doi: 10.1126/science.aaf9794.

